# Latent Tuberculosis Infection in Focus: Evaluating Public Concerns, Treatment Perceptions, and Awareness in King Sabatha Dalindyebo Local Municipality, Eastern Cape, South Africa

**DOI:** 10.1101/2025.08.12.25333508

**Authors:** Lindiwe Modest Faye, Ncomeka Sineke, Ntandazo Dlatu, Urgent Tsuro, Mojisola Clara Hosu, Teke Apalata

## Abstract

**Background:** Latent tuberculosis infection (LTBI) remains an overlooked aspect of tuberculosis (TB) control in South Africa, despite its potential to develop into active disease. This study aimed to assess public awareness, perceptions of treatment, and perceived barriers to LTBI care in a high-burden rural setting.

**Methods:** A cross-sectional, community-based survey was conducted among adults in the King Sabatha Dalindyebo Local Municipality, Eastern Cape. A structured questionnaire assessed knowledge of LTBI, concerns about progression, perceived consequences, and treatment-related barriers. Data were analyzed using descriptive statistics, chi-square tests, and Pearson correlation, with results presented by demographic subgroup.

**Results:** Awareness of LTBI varied significantly by age, with the highest awareness among 20–29-year-olds (52.7%; 95% CI: 44.7–60.7; *p* = 0.5625) and the lowest in those aged 50– 59 (15.8%; 95% CI: 5.5–37.6; *p* = 0.0044). Belief in the necessity of treatment was strongest in the <20 group (66.7%) and those aged 20–29 (62.3%; 95% CI: 54.2–69.8; *p* = 0.0036), though actual treatment uptake remained low across all age groups, peaking at 14.8% (95% CI: 5.9–32.5; *p* = 0.0003) in the 30–39-year-old cohort.

Concern about LTBI progressing to active TB was widespread (75.1%; 95% CI: 69.3–80.1; *p* < 0.0001), but only 38.3% correctly identified the full consequences of untreated LTBI (*p* = 0.0012). The most commonly cited barrier was lack of awareness (62.4%; 95% CI: 56.2–68.3; *p* < 0.0001). Correlation analysis revealed a weak but significant association between understanding of consequences and reduced informational barriers (*r* = 0.186; 95% CI: 0.064–0.307; *p* = 0.0035), while emotional concern was not significantly associated with actual knowledge or behavior.

**Conclusion:** The findings highlight a disconnect between concern and action in LTBI care. While awareness and worry are common, they are insufficiently grounded in accurate knowledge or translated into treatment uptake. Community education, stigma reduction, and age-sensitive messaging are crucial for enhancing LTBI engagement and supporting TB prevention in rural South Africa.

## Introduction

Tuberculosis (TB) remains a critical public health challenge in South Africa, a country that continues to bear one of the highest burdens of TB globally [1, 2, 3]. While much attention is paid to active TB, latent tuberculosis infection (LTBI) represents a silent yet significant component of the disease burden. LTBI refers to a state in which individuals are infected with *Mycobacterium tuberculosis* but do not exhibit clinical symptoms and are not infectious [4,5]. However, approximately 5–10% of individuals with LTBI progress to active TB in their lifetime, with this risk being markedly higher among those with immunosuppression, including HIV infection [4,6,7]. Despite the availability of diagnostic and preventive treatment strategies, LTBI remains poorly addressed in many high-burden, resource-limited settings. Challenges include limited awareness among the public and healthcare providers, stigma, fear of side effects, and barriers to accessing treatment [8,9,10]. In South Africa, these challenges are compounded by social inequalities, inadequate health infrastructure, and limited community engagement around preventive TB strategies [11,12].

This study focuses on assessing community-level concerns, awareness, and perceptions of treatment related to LTBI in the King Sabatha Dalindyebo (KSD) Local Municipality, Eastern Cape. By evaluating public understanding and identifying barriers to engagement with LTBI care, this research aims to inform context-specific interventions to improve LTBI diagnosis, treatment uptake, and TB prevention outcomes.

## Methodology

### Study design and setting

This was a cross-sectional, community-based survey conducted in the KSD Local Municipality, situated in the Oliver Reginald (O.R.) Tambo District of the Eastern Cape Province. The area is characterized by a high burden of TB and HIV, limited healthcare access in rural settlements, and persistent social inequalities affecting health-seeking behavior.

### Sampling and participants

Participants were recruited using random sampling from various communities in the municipality and clinics across the municipality. The inclusion criteria were individuals aged 18 years and above, residing in the KSD Local Municipality, and able to provide informed consent.

### Data collection tool

Table 1 outlines the structured questionnaire items administered to participants in the KSD Local Municipality. Questions assessed awareness of LTBI, perceived consequences, emotional concern, treatment beliefs, testing history, and barriers to care. The tool was adapted from World Health Organization (WHO) TB education guidelines, the tool covered key domains including demographic characteristics, awareness and understanding of LTBI (e.g., Q1, Q6), attitudes toward LTBI treatment (Q11–Q13), perceived barriers to testing and treatment (Q14), and history of LTBI screening or treatment (Q15–Q19) (Table 1). To ensure cultural relevance and clarity, the questionnaire was translated into isiXhosa for linguistic and contextual appropriateness.

### Data analysis

Descriptive statistics were used to summarize demographic variables and LTBI-related knowledge and attitudes. Proportions, 95% confidence intervals (CIs), and p-values were calculated to assess significant differences across age, gender, and ethnicity subgroups. Binary variables were generated for multivariate analysis and correlation testing. Pearson’s correlation coefficients (r), 95% CIs, and p-values were computed to evaluate associations between concern, awareness, and perceived barriers. A network visualization was constructed using correlation weights to illustrate the strength of relationships among key constructs.

### Ethical considerations

The study was conducted following the Declaration of Helsinki and approved by the Research Ethics and Biosafety Committee of the Faculty of Medicine and Health Sciences of Walter Sisulu University (ref. no. 084/2024) and the Eastern Cape Department of Health (ref. No. EC_202409_008).

## Results

Table 2 (supp file 2) presents an analysis of awareness and treatment perceptions related to LTBI across demographic groups, including age, gender, and ethnicity. Awareness of LTBI varied notably across age groups. Out of 245 participants, the highest awareness was observed in participants aged 20–29 years (52.7%; 95% CI: 44.7–60.7), whereas individuals aged 60 and above had the lowest awareness (18.2%; 95% CI: 5.1–47.7). Although younger participants (<20 years) had moderate awareness (40.0%), their belief in treatment necessity and uptake was minimal. Gender and ethnicity trends followed similar patterns, though statistically significant differences were mainly seen in age-based comparisons.

In Table 3 (supp file 2), participants aged 20–29 years demonstrated the highest belief in the necessity of LTBI treatment (62.3%; 95% CI: 54.2–69.8; p = 0.0036), However, actual treatment uptake in this group remained low (6.2%), reflecting a disconnect between awareness and action. The 30–39-year group exhibited moderate belief (55.9%) but reported the highest treatment uptake (14.8%; 95% CI: 5.9–32.5). In contrast, the 40–49 and 60+ age groups reported relatively low belief in treatment (40.0% and 45.5%, respectively), with minimal or no uptake, particularly among those aged 60 and above, where uptake was 0%. Among participants under 20 years, belief in treatment was relatively high (66.7%), but no uptake was recorded. Gender-specific patterns, though not fully tabulated here, indicated that females generally reported stronger belief in treatment necessity and slightly higher treatment uptake. Ethnicity trends, primarily among Black South African respondents, the dominant demographic in this sample, mirrored broader findings of moderate to high treatment perception but consistently low uptake. Notably, the most frequently cited barrier to LTBI testing or treatment was a lack of awareness (62.4%), followed by fear of side effects (14.7%) and stigma associated with TB (13.9%). Financial constraints were reported less often but remain a potential obstacle.

Table 4 (supp file 2) summarizes key findings on public concerns, perceived consequences, and barriers to LTBI treatment. Approximately 75.1% of participants reported being either very or somewhat concerned about the risk of LTBI progressing to active tuberculosis (95% CI: 69.3–80.1; p < 0.0001. However, only 38.3% accurately identified the full range of consequences associated with untreated LTBI (95% CI: 31.7–45.2). The most prevalent barrier to testing and treatment was lack of awareness, cited by 62.4% of respondents (95% CI: 56.2–68.3; p < 0.0001).

Table 5 (supp file 2) presents Pearson correlation coefficients (r), confidence intervals (CI), and p-values for pairwise associations among community concerns about LTBI progression, understanding of its health consequences, and three commonly cited barriers to testing or treatment (lack of awareness, fear of side effects, and stigma). Statistically significant associations are highlighted, with the strongest positive correlation observed between perceived consequences and lack of awareness (r = 0.186, p = 0.0035), suggesting that improved understanding may reduce informational barriers.

Fig.1 (Supp file 1) is a correlation analysis conducted to explore the relationships among concern about LTBI progression, perceived consequences of untreated infection, and the three most frequently cited barriers to testing or treatment. The only statistically significant correlation was observed between perceived consequences and lack of awareness as a barrier (r = 0.186; 95% CI: 0.064–0.307; p = 0.0035). No statistically significant associations were found between concern about progression and any of the other variables, including perceived consequences (r = -0.089; p = 0.1668), lack of awareness (r = 0.119; p = 0.0634), fear of side effects (r = -0.054; p = 0.3974), or stigma (r = 0.040; p = 0.5331).

**Fig 1.**
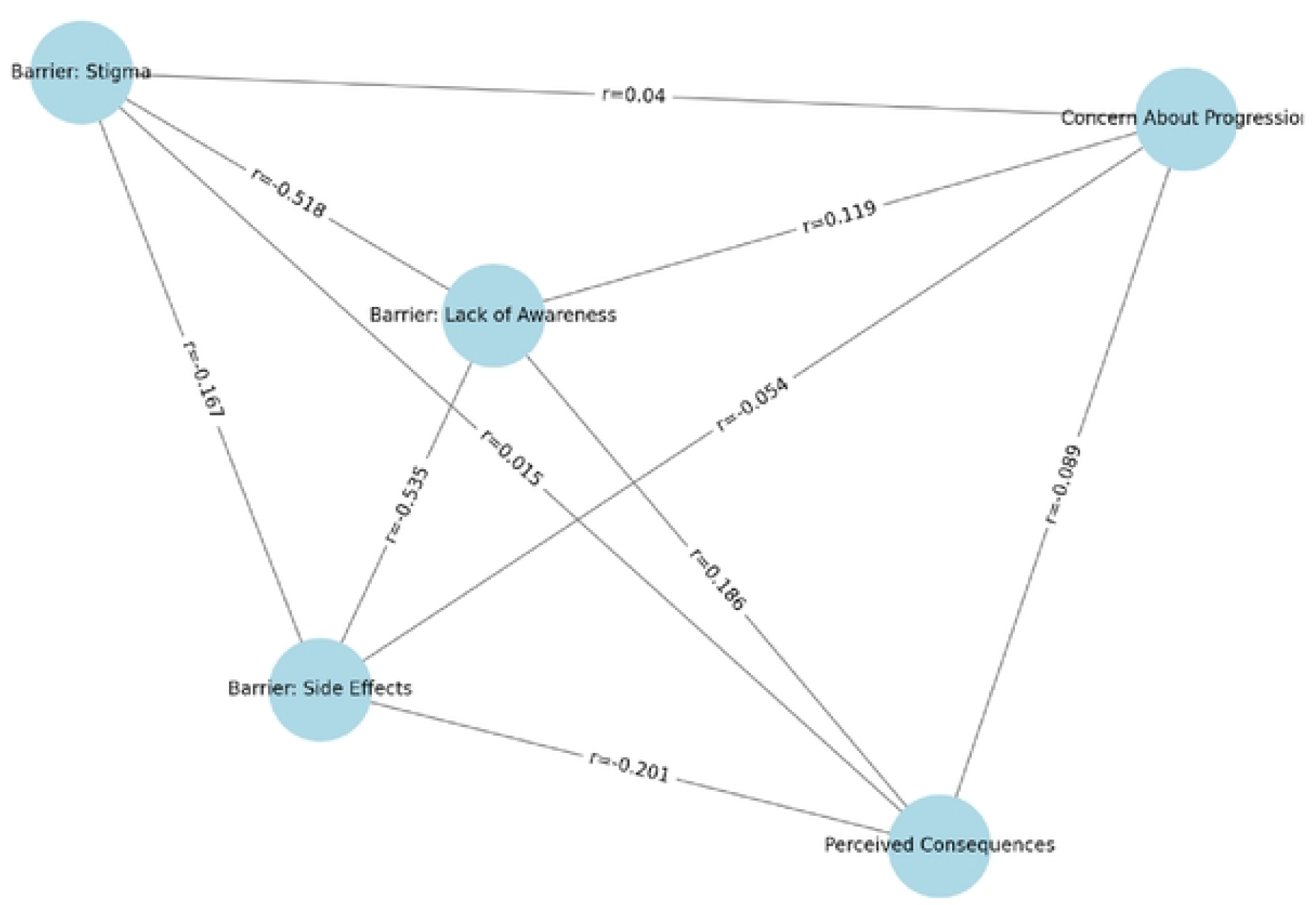
Correlation analysis ofLTBI perceptions and barriers.

The Predictive pathway of LTBI Intervention Impact in Figure 2 (Supp file 1) illustrates the theorized pathway from the current community-level challenges in the KSD Local Municipality to potential improvements in LTBI care and TB prevention. On the left, the model highlights key obstacles to LTBI treatment uptake, including low awareness, high concern with limited understanding, and persistent barriers such as stigma, fear of side effects, and restricted access to healthcare. These factors collectively contribute to poor engagement with LTBI screening and preventive therapy. At the center, three targeted interventions community education campaigns, stigma reduction through community health worker (CHW) involvement, and age- and gender-sensitive health messaging are proposed to directly address these root causes. The right side of the model projects the anticipated outcomes if such interventions are implemented: enhanced community knowledge of LTBI, reduced social and psychological barriers, increased uptake of treatment, and ultimately, improved TB prevention and quality of life in the affected population.

**Fig 2.**
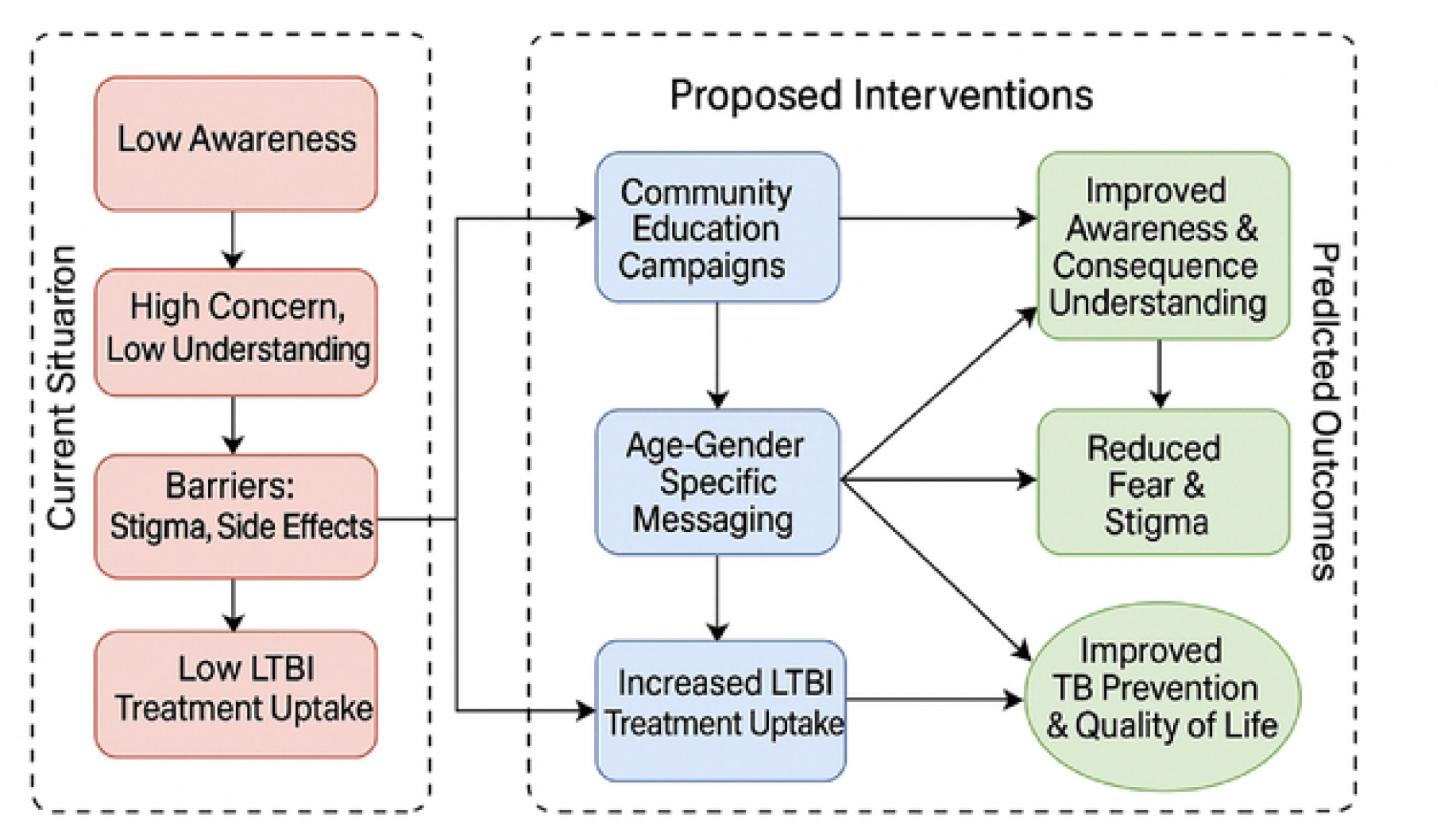
Predictive Pathway for improved LTBI outcomes.

## Discussion

This study highlights a complex and often fragmented understanding of LTBI among community members in the KSD Local Municipality. While general awareness of LTBI was moderate, and concern about disease progression was high, these did not consistently translate into an accurate understanding of consequences or meaningful engagement with testing and treatment. Participants aged 20–29 exhibited the highest belief in the necessity of LTBI treatment, while those aged 30–39 had the highest actual uptake, although both rates remained low [13,14]. Various studies on LTBI demonstrated different patterns of belief in the need for treatment and how often people start it across various age groups. For example, while younger adults may see LTBI treatment as more necessary, actual participation can be affected by a complex mix of factors, with other age groups possibly showing higher completion rates because of different motivations or fewer obstacles [15,16,17]. These findings suggest a gap between intention and behavior regarding LTBI treatment, likely driven by a combination of knowledge gaps, social stigma, and systemic barriers [18,16,9]. This is further supported by the strong identification of lack of awareness (62.4%) as a leading barrier, followed by fear of side effects (14.7%) and TB-related stigma (13.9%). Such specific findings align with broader literature emphasizing these as significant impediments to LTBI care [19,15]. Correlation and network analyses reinforced this disjunction. While concern about progression was high (75.1%), it was not significantly associated with accurate knowledge or barrier-related perceptions [20]. The only statistically significant correlation observed was between perceived consequences and lack of awareness (r = 0.186, p = 0.0035), suggesting that improved understanding may reduce informational barriers [17, 21]. The study by Magwaza et al. also highlights similarities in the role of “ever heard of LTBI before” and its negative correlation with understanding progression, which is consistent with our findings regarding “lack of awareness” and “perceived consequences [17].” These findings underscore the need for integrated community education campaigns that move beyond raising general awareness to addressing the specific misconceptions and fears associated with LTBI. Tailored messaging, stigma reduction interventions, and community health worker engagement may be necessary to close the gap between perception and action. Furthermore, interventions should be stratified by age and gender to ensure context-specific relevance, with particular focus on empowering younger adults and supporting older, at-risk individuals.

The correlation network visually represents the relationships among community concern about LTBI progression, understanding of its health consequences, and perceived barriers to testing or treatment. Nodes represent key constructs, concerns, perceived consequences, and three prominent barriers, while edges indicate the strength and direction of their statistical association (based on Pearson’s correlation coefficients).

The most notable connection in the network is between Perceived Consequences and Barrier is Lack of Awareness (r = 0.186), the only statistically significant correlation (p = 0.0035) [17]. This edge suggests that individuals who have a better understanding of the consequences of untreated LTBI are also more likely to be aware of LTBI itself, highlighting the foundational role of comprehensive health knowledge in overcoming informational barriers. These findings are consistent with similar results in Canada, among Chinese immigrants, and in three Southeastern counties in the United States ^[22,23]^. Conversely, the central node’s concern about progression is only weakly connected to other variables in the network. Its low or nonsignificant correlations with perceived consequences, fear of side effects, and stigma (all p > 0.05) suggest that emotional concern about disease progression may exist independently of factual understanding or perceived structural barriers ^[24]^. This separation implies that concern may be driven by generalized fear or hearsay rather than informed risk assessment. Additionally, the lack of significant association between concern and stigma contradicts assumptions that fear of TB-related discrimination directly influences anxiety about disease progression. Similarly, the weak connection between concern and side effects indicates that worry about progression does not strongly predict fear of treatment harms [25,26]. Besides perceptions and uptake, this study evaluated public concern regarding LTBI progression, understanding of its consequences, and barriers to testing or treatment. A significant proportion of respondents expressed themselves as being either very concerned or somewhat concerned about progression from LTBI to active TB. This indicates a general awareness of TB risks. When asked about the consequences of untreated LTBI, most participants correctly identified serious health implications, including disease progression and transmission. However, this concern and understanding did not necessarily translate into treatment uptake.

The predictive model reinforces findings from this study, highlighting a significant disconnect between concern and action regarding LTBI care. While participants express anxiety about disease progression, this concern is not grounded in factual understanding or accompanied by proactive behavior, such as testing or treatment uptake. This disconnect between perceived risk and preventive behavior, such as testing, treatment initiation, or adherence, is both revealing and consistent with findings from other studies. A similar behavioral disconnect was observed by Alsdurf et al., who described a cascade of care model for LTBI and noted significant attrition at each stage from testing to treatment completion. They attributed this to low health literacy, poor risk perception, and systemic barriers, particularly in vulnerable populations. Despite the worry about progression to active TB, patients failed to act, often due to inadequate understanding of LTBI and its management [27]. In a South African context, a study by Louw reported that community members who feared TB infection often delayed testing and treatment because their concerns were not rooted in accurate knowledge [28]. Their study underscored that misconceptions and low perceived personal risk contributed to reduced uptake of preventive therapy, even among household contacts of active TB patients. Contrastingly, a study conducted in Ethiopia found that TB-related knowledge strongly predicted testing behavior. In communities with targeted education campaigns, individuals were more likely to take preventive action when they understood LTBI transmission and treatment implications. This suggests that contextual and educational interventions can realign concern with evidence-based action. Moreover, Zenner et al. emphasized in their UK-based LTBI screening program that psychological readiness and informed consent were crucial for high uptake. Participants who received tailored health information were more willing to participate in testing and complete treatment. Together, these studies illustrate that while emotional concern may exist, actionable engagement hinges on an accurate understanding and access to supportive services. This reinforces the need for interventions that do more than raise alarm but must empower individuals with practical knowledge and resources [29].

The model suggests that community-level education is essential to convert emotional concern into informed decision-making. This behavioral inertia can largely be attributed to gaps in health literacy, especially in resource-limited and rural settings. Here, community-level education becomes an essential lever for change. Community-based education can close knowledge gaps by explaining LTBI risk, treatment options, and outcomes [30]; Address stigma and misconceptions, especially in rural settings where TB is often misunderstood; Promote agency, empowering individuals to act on their concerns and enhance healthcare trust, particularly when delivered by community health workers. In Ethiopia, Datiko et al, found that empowering health extension workers to engage directly with community members increased awareness and testing uptake [31]. Additionally, integrating community health workers and reducing LTBI- and TB-related stigma are likely to enhance trust and access, particularly for women, youth, and older adults who face different sets of challenges [32].

Tailored communication is especially relevant, as the study revealed age-specific patterns: younger respondents were more concerned but less likely to act, while those aged 30–39 had higher follow-through rates. This suggests that uniform strategies may be ineffective without considering demographic context [33]. The projected endpoint, which improves quality of life and TB prevention, aligns with South Africa’s national TB control priorities and the WHO End TB Strategy. The model provides a foundation for operationalizing study insights into practical, scalable interventions that can bridge the gap between awareness and preventive action in high-burden, rural settings. In summary, the network reveals that while concern is widespread, it is not well integrated with accurate knowledge or perceptions of barriers. This disjunction underscores the need for interventions that simultaneously improve LTBI literacy, reduce informational barriers, and ensure concerns are grounded in accurate health understanding.

## Recommendations

Based on the findings of this study, several targeted strategies are recommended to strengthen LTBI care in rural, high-burden settings such as the KSD Local Municipality. First, community-based education campaigns should be prioritized to improve awareness of LTBI, its potential progression, and the importance of treatment even in asymptomatic individuals. These messages must be tailored by age and gender to address the differing perceptions and uptake behaviors identified across demographic groups. Second, stigma reduction initiatives should be integrated into TB outreach, using trusted CHWs to build local trust and dispel misconceptions around LTBI testing and treatment. Third, policy-level support is needed to ensure the inclusion of LTBI education in existing TB prevention programs and to remove logistical barriers such as limited-service availability or transportation challenges. Lastly, further research should explore health system readiness to deliver LTBI preventive therapy at scale and investigate the role of digital platforms in reinforcing community engagement.

## Conclusions

This study highlights a significant gap between public concern and action regarding LTBI in a rural South African setting. While many community members worry about LTBI developing into active TB, this concern is not backed by sufficient knowledge, and it does not lead to high treatment rates. Low awareness, misconceptions about the consequences, and persistent social barriers such as stigma and fear of side effects continue to hinder engagement with LTBI services. Correlation analysis shows that accurate knowledge is a better predictor of reducing barriers than concern alone, emphasizing the importance of informed health literacy as a catalyst for behavior change. Addressing these gaps through culturally appropriate, community-led interventions will be crucial for improving TB prevention efforts and health outcomes among vulnerable populations.

## Author Contributions

Conceptualization, L.M.F.; N.D.; and N.S.; methodology, L.M.F.; N.S.; validation, L.M.F., and N.S.; formal analysis, L.M.F.; U.T.; investigation, L.M.F.; N.D.; and M.C.H.; resources, L.M.F.; data curation, U.T.; writing—original draft preparation, N.S.; L.M.F.; writing—review and editing, M.C.H., N.D. and T.A.; visualization, L.M.F.; U.T.; supervision, T.A.; project administration, L.M.F. All authors have read and agreed to the published version of the manuscript.

## Informed Consent Statement

This is not applicable as this study only reviewed patient files.

## Data Availability Statement

Data can be requested from the corresponding author.

## Acknowledgments

We are grateful for student support during data collection, Mr. Cebo Magwaza.

## Supplementary File 1 Figures

